# Hepatic Transthyretin knockdown alleviates NAFLD by enhancing SERCA2 function and inhibiting endoplasmic reticulum stress

**DOI:** 10.1101/2025.10.15.25338095

**Authors:** Yingzi He, Tian Yang, Ruojun Qiu, Bingyang Liu, Shuo Wang, Jianan Wang, Fenping Zheng

**Author notes:** Address correspondence to: Fenping Zheng, Department of Endocrinology, the Affiliated Sir Run Run Shaw Hospital, College of Medicine, Zhejiang University, Hangzhou, Zhejiang, 310016, China.

## Abstract

Endoplasmic reticulum (ER) stress is considered a key trigger in the nonalcoholic fatty liver disease (NAFLD). Dysfunction of sarcoplasmic/endoplasmic reticulum calcium ATPase 2 (SERCA2) leads to insufficient ER Ca^2+^ storage, thus activating ER stress. Transthyretin (TTR), expressed and secreted by the liver, may function as a new “hepatokine” related with insulin resistance (IR). However, the role of TTR in NAFLD remains unclear. We firstly found the hepatic *Ttr* mRNA expression was elevated in NAFLD patients. TTR over-expression exacerbated steatosis in fatty livers, while TTR knockdown alleviated IR, steatosis, inflammation, and upregulated SERCA2 expression, leading to reduced ER stress in NAFLD mice. Consistently, TTR knockdown alleviated lipid deposition in steatotic hepatocytes. TTR stimulation of hepatocytes increased cytosolic Ca²⁺ and decreased ER Ca²⁺. Conversely, in steatotic AML12 cells, TTR knockdown increased ER calcium storage and mitigated the cytosolic calcium pulse induced by thapsigargin. Then using immunofluorescence and co-immunoprecipitation, we further confirmed the interaction between TTR and SERCA2. In conclusion, TTR knockdown ameliorated liver steatosis and inflammation in NAFLD, which might be related to the improvement of cellular calcium homeostasis and ER stress by reversing SERCA2 function.

## Introduction

Nonalcoholic fatty liver disease (NAFLD) is a continuous spectrum that includes simple non-alcoholic fatty liver (NAFL) and non-alcoholic steatohepatitis (NASH), with a portion progressing to cirrhosis and even hepatocellular carcinoma. NAFLD is a chronic metabolic disease closely related to obesity and metabolic syndrome(***Quek et al., 2023***). With an “obesity epidemic” raging throughout many parts of the world, NAFLD has become the most common chronic liver disease worldwide(***Henry et al., 2022*; *Paik et al., 2020***). However, the pathogenesis of this disease is complex and largely unknown. The “multiple hits theory” indicates that factors such as insulin resistance (IR), mitochondrial dysfunction, endoplasmic reticulum (ER) stress, changes in gut microbiota, genetics, and epigenetics are all involved(***Buzzetti et al., 2016*; *Loomba et al., 2021***). The complex pathogenesis poses a significant challenge for discovering effective treatments for the disease.

Excessive and sustained ER stress is considered to be a key trigger in the progression of NAFLD(***Rutkowski et al., 2008***). ER is the primary sub-cellular organelle for lipid synthesis and calcium store. The storage of calcium by ER is the most critical process for maintaining the intra-cellular calcium homeostasis. In recent years, the impact of calcium signaling on NAFLD, particularly in terms of lipid accumulation, has garnered significant attention, and targeting the regulation of calcium homeostasis emerges as a novel intervention strategy for combating NAFLD(***Park and Lee, 2014***). The sarcoplasmic/endoplasmic reticulum calcium ATPase (SERCA) is the sole calcium uptake channel and is essential for maintaining the high concentration of Ca^2+^ in the ER(***Periasamy and Kalyanasundaram, 2007; Vangheluwe et al., 2005***). Dysfunction of SERCA leads to insufficient Ca^2+^ storage in the ER, resulting in protein misfolding and subsequently activating ER stress and thus altering lipid synthesis(***Krebs et al., 2015***). SERCA2 is the primary isoform of SERCA in the liver(***Vangheluwe et al., 2005***). Plenty of studies have demonstrated that enhancing the function of SERCA2 can ameliorate ER stress and delay the progression of NAFLD(***Jung et al., 2018; Gao et al., 2018; Park et al., 2010; Kang et al., 2016***). Therefore, targeting the modulation of SERCA function to improve ER calcium homeostasis may offer a novel strategy for the treatment of NAFLD.

Vast researches indicate that liver is capable of expressing a variety of genes that encode extracellular proteins, termed “hepatokines”, which participate in the metabolic regulation by establishing inter-organizational “cross-talk” through autocrine, paracrine, and endocrine actions(***Weigert et al., 2019; Iroz et al., 2015***). Our studies have shown that transthyretin (TTR) may function as a “hepatokine”. In different mouse models of IR, hepatic TTR expression and secretion are increased. Excessive TTR induces the occurrence of IR and inhibit the insulin-sensitizing effect of exercise in obese mice(***He et al., 2021***). Another study also showed that TTR is directly involved in the regulation of IR and glucose metabolism(***Zemany et al., 2015***). Additionally, its impact on calcium homeostasis is also noteworthy. TTR causes Ca^2+^ oscillations in pancreatic cells(***Dekki et al., 2012***) and in skeletal muscle cells, as our previous findings reported(***Wu et al., 2023***). However, the association between TTR and the development of NAFLD is still unclear. Given the pivotal role of IR and calcium homeostasis in the development of NAFLD, we hypothesize that TTR may be involved in the onset and progression of NAFLD.

## Results

### Expression of TTR is increased in patients with NASH and TTR over-expression causes hepatic lipid deposition in vitro and in vivo

In our previous study, we found increased hepatic *Ttr* expression and secretion in patients with metabolic syndrome and in different obese mouse models(***He et al., 2021***). NAFLD is also considered a state of IR, and a complication of obesity and metabolic syndrome, so we measured *Ttr* mRNA expression in the liver biopsy specimen of patients with NASH. Figure 1A demonstrated that the patients with NASH exhibited higher hepatic *Ttr* mRNA expression compared with the non-NAFLD individuals. Then we constructed mouse models with TTR over-expression by tail vein injection of TTR-expressing adenovirus (Ad-*Ttr*) into the mice after 8 weeks of high-fat-diet (HFD) feeding. The hepatic TTR expression in mice fed on HFD or normal diet (ND) was increased after injection with Ad-*Ttr* (Figure 1B), as was the increased serum TTR levels (Figure 1C). TTR over-expression didn’t affect liver lipid contents in ND mice, but further aggravated hepatic steatosis in HFD mice (Figure 1D, E). In *vitro*, both 0.75 mM of free fatty acids (FFAs) and 400 µM of palmitic acid (PAs) elevated *Ttr* mRNA expression in AML12 cells (Supplementary Figure 1). We consistently found that TTR up-regulation, achieved by *Ttr*-adenovirus infection exacerbated FFAs-induced lipid deposition in AML12 cells (Figure 1F-H). Thus, we firstly confirmed that excessive TTR expression under IR state in the liver leads to lipid deposition within it.

**Figure 1.**
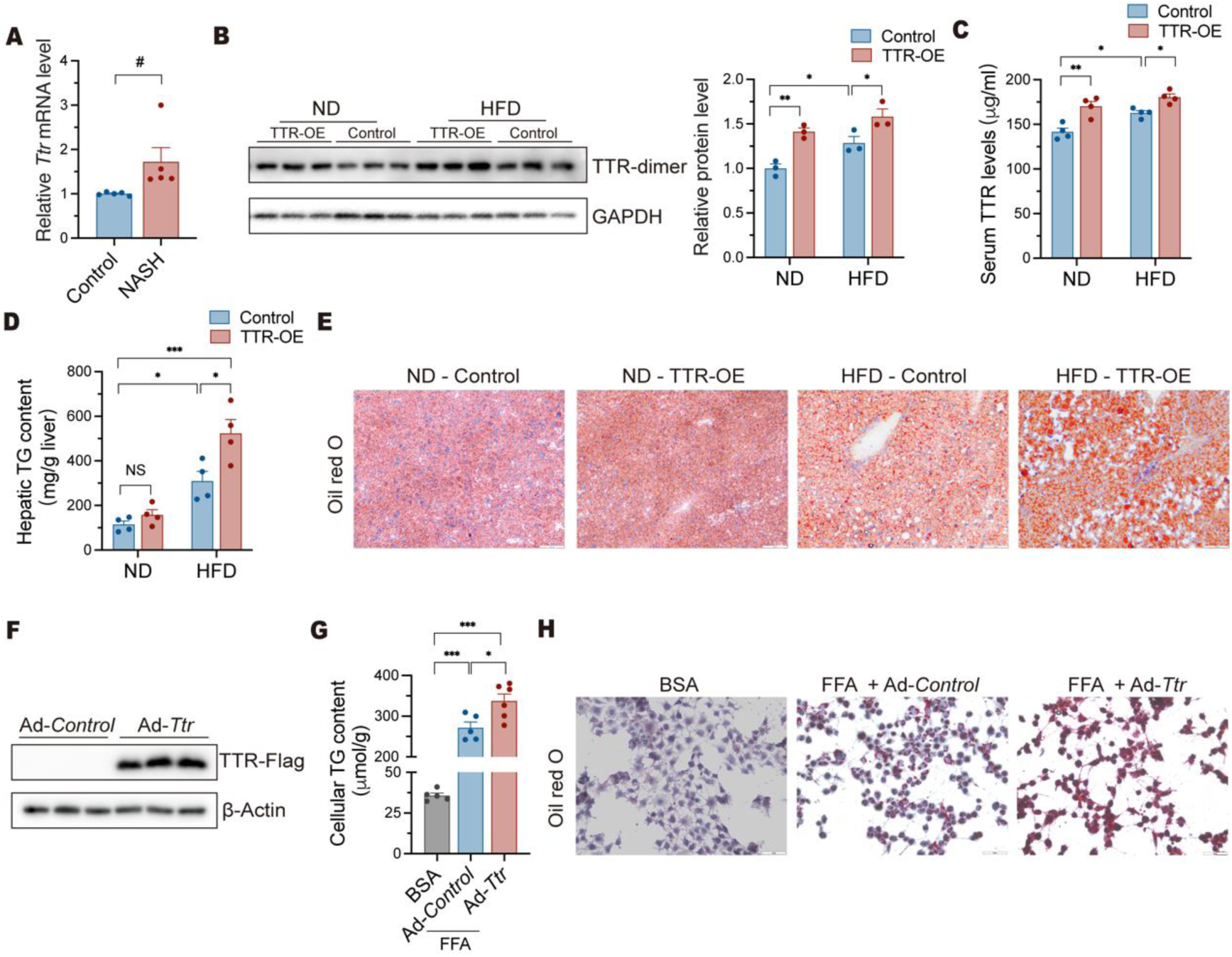
Elevated TTR is associated with the development of NAFLD. (A) Quantitative real-time PCR (qRT-PCR) detected *Ttr* mRNA levels from liver sections in NASH patients and non-NAFLD controls (n = 5 per group). (B-E) TTR was overexpressed in ND or HFD-fed mice by tail vein injection of adenovirus (n = 4 per group). (B) Western blot analysis of hepatic protein level of TTR. (C) Serum TTR levels were detected by ELISA. Hepatic TG contents (D) and Oil Red O staining (E) in mouse liver tissues. Scale bar: 100 μm. (F-H) TTR was overexpressed in AML12 cells by adenovirus. (F) Western blot analysis of protein level of TTR-FLAG. Hepatic TG contents (G) and Oil Red O staining (H) in AML12 cells. Scale bar: 100 μm. Data are shown as the mean ± SEM. *P* values were from 2-tail unpaired *t* test (A) and 1-way ANOVA (B, C, D and G). ^#^*P* = 0.057, \**P* < 0.05, \*\**P* < 0.01, \*\*\**P* < 0.001. ND, normal diet; HFD, high fat diet; Ad-*Ttr*, Adenovirus-*Ttr*; NS, no significance; TTR-OE, TTR overexpression; TG, triglyceride.

### Liver-specific TTR knockdown (TTR-KD) improved glucose tolerance and insulin tolerance in HFD and Gubra-Amylin NASH (GAN)-diet mice

To further investigate whether TTR plays a role in the development of NAFLD, we established HFD/GAN diet-induced mouse models with liver-specific TTR knockdown by tail vein injection of adeno-associated virus-sh*Ttr* bearing TBG promoter (AAV-TBG-sh*Ttr*). After 8 weeks of AAV injection into HFD or GAN-diet mice, the hepatic expression and secretion of TTR into circulation decreased compared with the control groups (Figure 2A, B, C, G, H, I). Specific TTR knockdown in liver didn’t decrease the body weight of mice fed on 19 weeks of HFD or 17 weeks of GAN diet (Figure 2D, J) and the food intake of mice fed with GAN diets (Supplementary Figure 2), which was comparable to mice fed on ND. However, glucose tolerance and insulin sensitivity were significantly improved in mice fed on HFD or GAN diet by TTR knockdown (Figures 2E, K, F, L).

**Figure 2.**
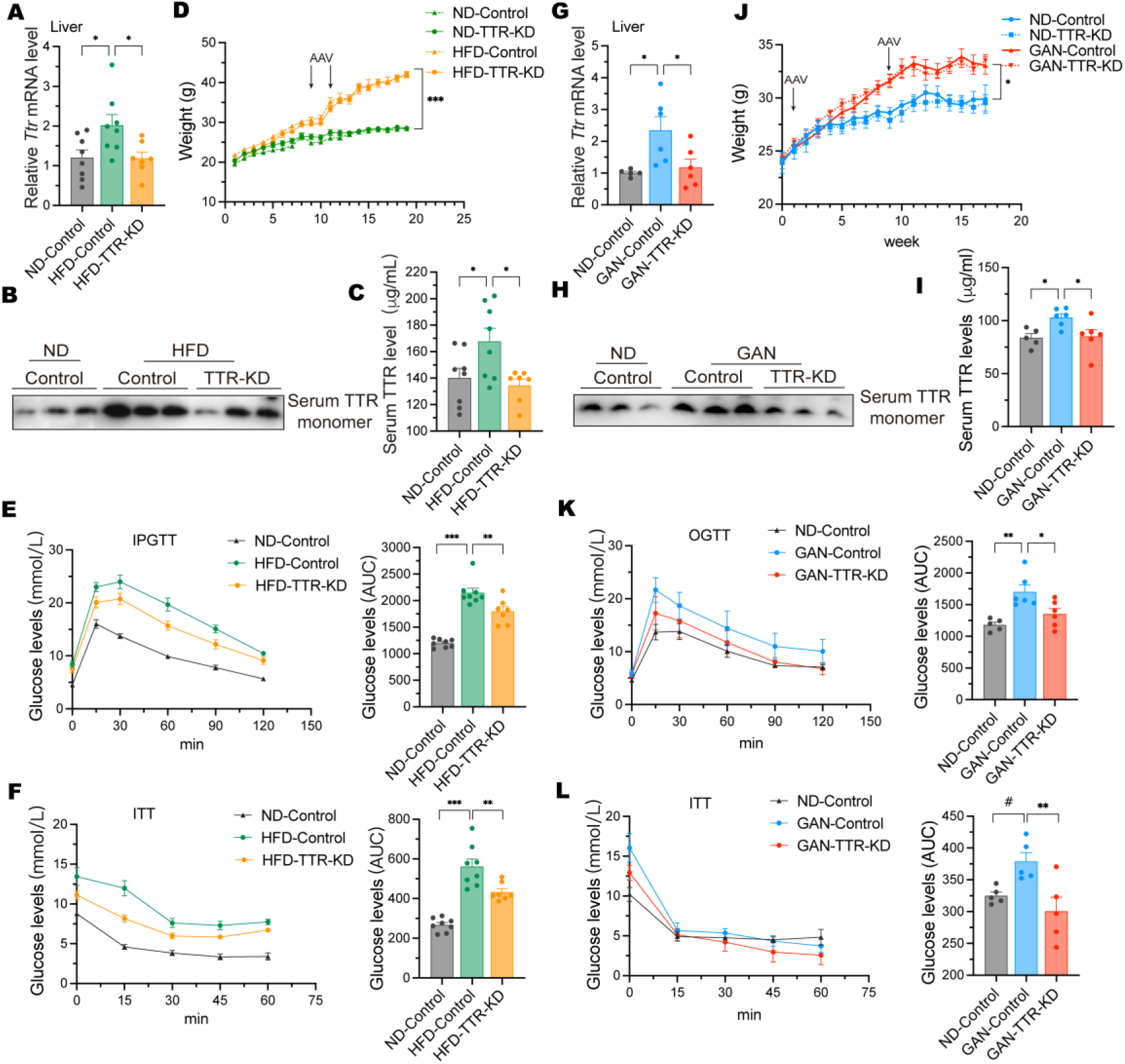
The effect of TTR-KD on insulin resistance. (A-F) TTR knockdown was achieved in the HFD-fed mice as mentioned in the Materials and Methods section (n = 7-8 per group). (A) qRT-PCR detected hepatic *Ttr* mRNA levels in HFD-fed mice. Serum TTR levels were examined by western blot (B) and ELISA (C). (D) The body weight of HFD-fed mice was recorded per week for 19 weeks. Blood glucose levels and AUC of IPGTT (E) and ITT (F) in HFD-fed mice. (G-L) TTR knockdown was achieved in the GAN diet-fed mice as described in the Materials and Methods section (n = 5–6 per group). (G) qRT-PCR detected hepatic *Ttr* mRNA levels in GAN diet-fed mice. Serum TTR levels were measured by western blot (H) and ELISA (I). (J) The body weight of GAN diet-fed mice was recorded per week for 17 weeks. Blood glucose levels and AUC of OGTT (K) and ITT (L) in GAN diet-fed mice. Data are shown as the mean ± SEM. *P* values were from 1-way ANOVA (A, C, D, J, E, F, G, I, K and L). ^#^*P* = 0.07, \**P* < 0.05, \*\**P* < 0.01, \*\*\**P* < 0.001. TTR-KD, TTR knockdown; GAN, Gubra-Amylin NASH; IPGTT, intraperitoneal glucose tolerance test; ITT, insulin tolerance test; OGTT, oral glucose tolerance test; AUC, area under curve.

### Liver-specific TTR knockdown improved hepatic steatosis, inflammation, serum lipid and transaminase levels in HFD and GAN-diet mice

An increase in liver weight is an early symptom of fatty liver. We found that the liver weight (corrected by body weight) was significantly decreased in HFD mice by TTR knockdown (Figure 3A). Regarding to GAN-diet mice, the representative liver of TTR-KD mice was less greasy and redder than that of the control mice, although there was no difference in their liver weight (Figure 4 A, B). We further found that the liver TG contents, as well as lipid droplets in both HFD and GAN-diet mice, were decreased by TTR knockdown (Figure 3B, C) (Figure 4C, D).

**Figure 3.**
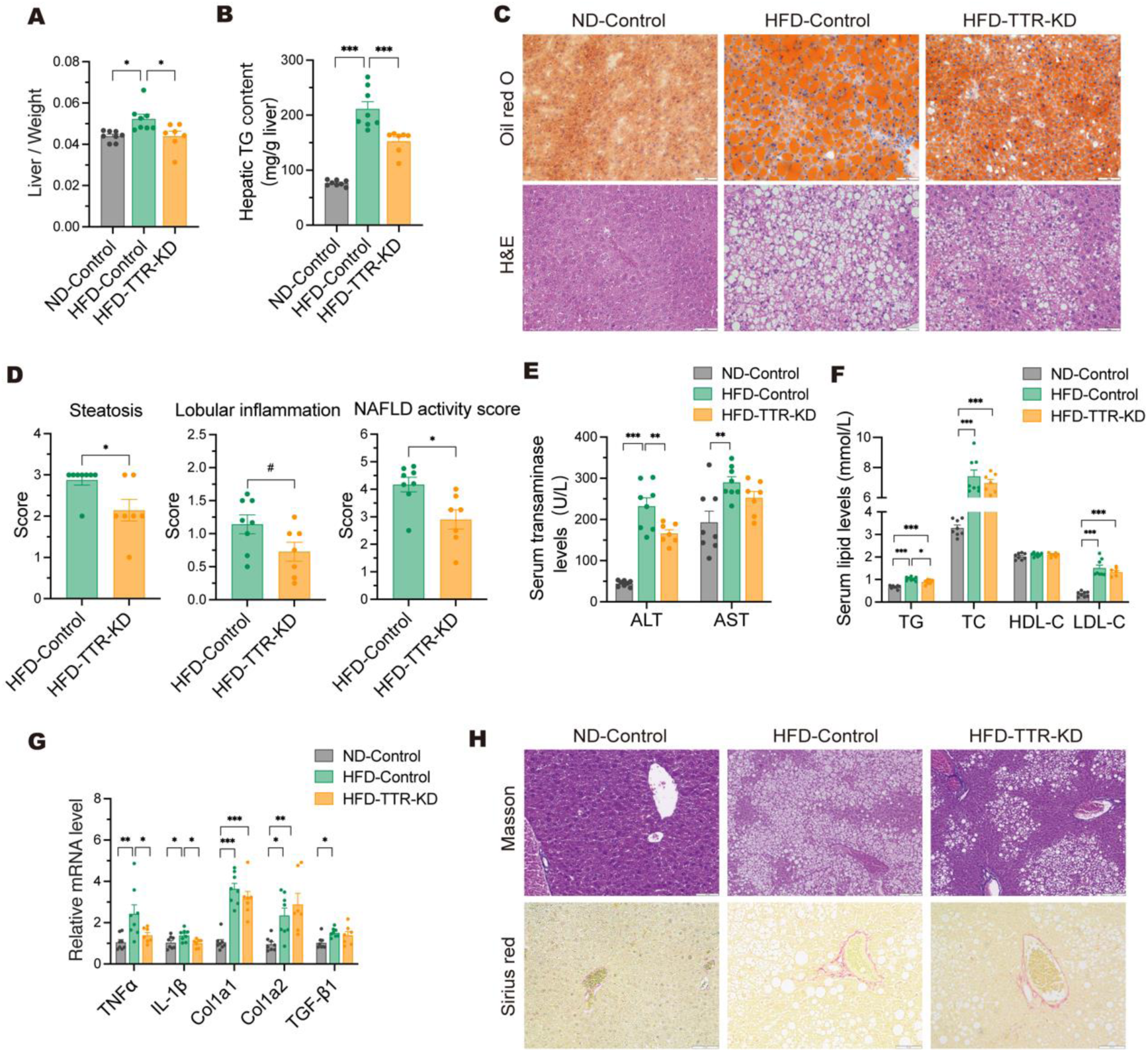
TTR knockdown improved liver lipid deposition, liver function as well as inflammatory gene expression in HFD-fed mice. Liver/body weight ratio (A) and hepatic TG contents (B) were measured in the TTR-KD and control mice. (C) Representative images of Oil Red O and H&E staining of liver tissue; scale bar: 100 μm. (D) Liver steatosis, lobular inflammation and NAFLD activity scores in HFD-fed mice. Liver function evaluated by serum ALT, AST levels (E) and lipid levels (F). (G) Related gene expressions of liver inflammation and fibrosis. (H) Representative images of Masson’s trichome and Sirius red staining of liver sections; scale bar: 100 μm. n = 7-8 per group. Data are shown as the mean ± SEM. *P* values were from 2-tail unpaired *t* test (D) and 1-way ANOVA (A, B, E, F and G). ^#^*P* = 0.06, \**P* < 0.05, \*\**P* < 0.01, \*\*\**P* < 0.001. H&E, hematoxylin and eosin; ALT, alanine aminotransferase; AST, aspartate aminotransferase; TC, total cholesterol; HDL-C, high-density lipoprotein cholesterol; LDL-C, low-density lipoprotein cholesterol.

**Figure 4.**
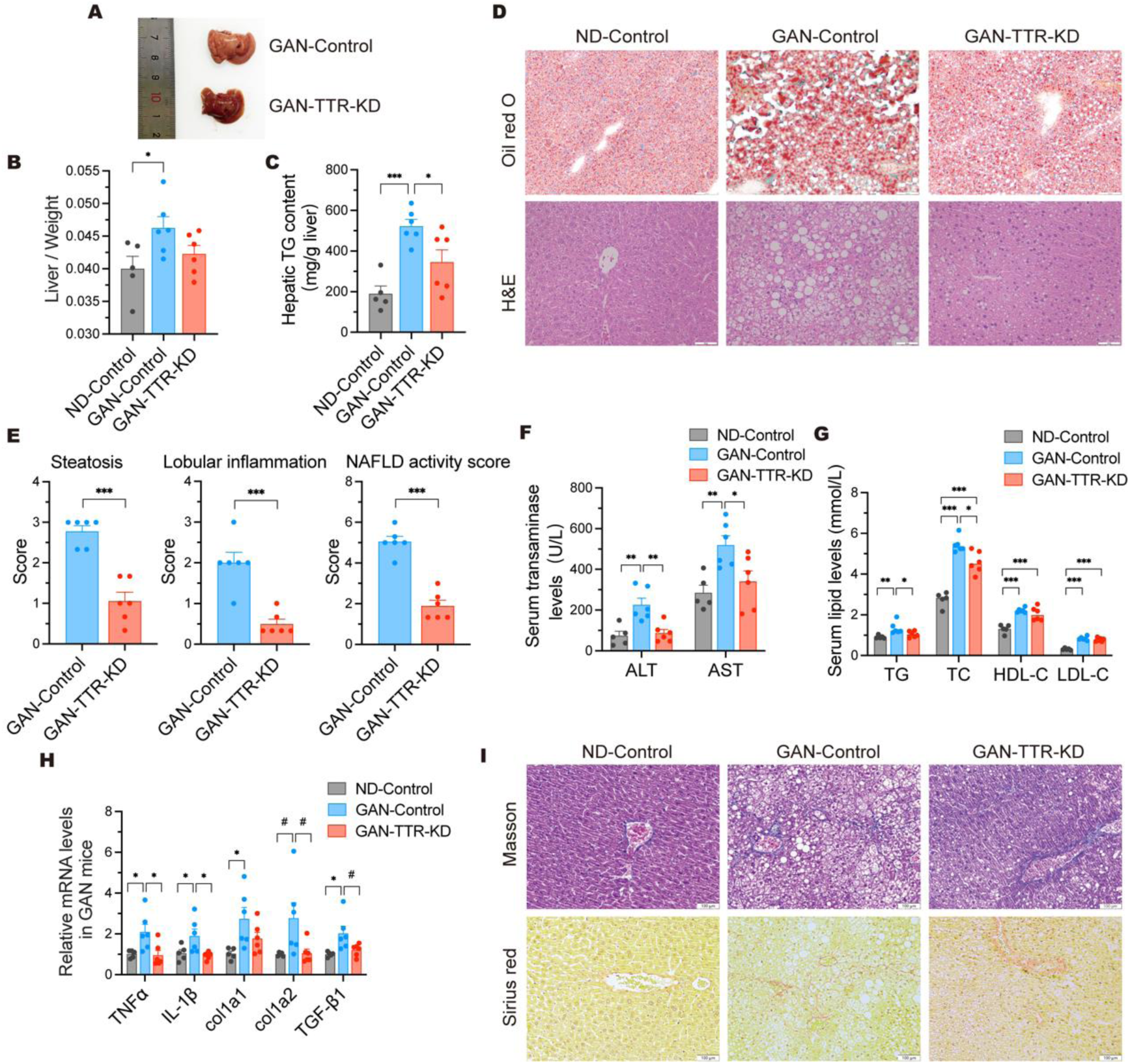
TTR knockdown improved liver lipid deposition, liver function as well as inflammatory gene expression, but did not significantly alter liver tissue fibrosis in GAN diet-fed mice. Representative images of general liver appearance (A), liver/body weight ratio (B) and hepatic TG contents (C) in the TTR-KD and control mice, respectively. (D) Representative images of Oil Red O and H&E staining of liver tissue; scale bar: 100 μm. (E) Liver steatosis, lobular inflammation and NAFLD activity scores in GAN diet-fed mice. Liver function evaluated by serum ALT, AST levels (F) and lipid levels (G). (H) Related gene expressions of liver inflammation and fibrosis. (I) Representative images of Masson and Sirius Red staining of liver sections; scale bar: 100 μm. n = 5-6 per group. Data are shown as the mean ± SEM. *P* values were from 2-tail unpaired *t* test (E) and 1-way ANOVA (B, C, F, G and H). ^#^*P* = 0.06, \**P* < 0.05, \*\**P* < 0.01, \*\*\**P* < 0.001.

The NAFLD activity score (NAS) is widely used for evaluating the severity of NAFLD, including steatosis, lobular inflammation and ballooning degeneration(***Kleiner et al., 2005***). We observed no obvious hepatocyte ballooning in the livers of mice fed on HFD or GAN diet, with an average NAS of 5.17 (Figure 3D) or 5.05 (Figure 4E), respectively. Yet, TTR knockdown significantly decreased the NAS (Figure 3D, 4E), alanine aminotransferase levels (Figure 3E, 4F), lipid levels (Figure 3F, 4G), and the gene expressions of inflammatory factors of *TNF-α* and *IL-1β* (Figure 3G, 4H), indicating the improvement of hepatic steatosis and inflammation in both mice. Early collagen deposition localizes to zone 3 of the hepatic lobule, showing perisinusoidal fibrosis with delicate blue fibers on Masson stain(***Gallage et al., 2022***). While TTR knockdown reduced the expression of fibrotic genes (*Col1a1*, *Col1a2*, and *TGF-β 1*) and decreased collagen deposition (assessed by Masson’s trichrome and Sirius red staining) in GAN-diet-fed mice (Figure 4H, I), but not in HFD-fed mice (Figure 3G, H).

Consistently, effectively knockdown of TTR expression in FFAs-induced AML12 cells by *Ttr* siRNA transfection (Supplementary Figure 3A, B) decreased the positive area in Nile red, as well as intra-cellular TG levels (Supplementary Figure 3C, D).

### TTR overexpression or knockdown did not influence mouse thyroid function

TTR plays a critical role as a primary thyroid hormone transporter in rodents. We thus evaluated the impact of TTR knockdown on circulating thyroid hormone levels. We found that circulating serum free thyroxine (FT4) levels were not affected by TTR knockdown (Supplementary Figure 4), indicating that TTR’s effect on IR or NAFLD was unlikely related to thyroid hormones.

### Liver-specific TTR knockdown affects lipid synthesis at ER levels in mice with NAFLD

TTR knockdown consistently ameliorated hepatic lipid accumulation across different NAFLD mouse models and in steatotic hepatocytes. To further explore the molecular mechanisms of improved NAFLD by TTR knockdown, we conducted transcriptomic analysis and non-targeted lipidomics analysis of liver samples from HFD mice. The Kyoto Encyclopedia of Genes and Genomes (KEGG) pathway analysis revealed that the differentially expressed genes (DEGs) were primarily enriched in glycerolipid metabolism and lipid and atherosclerosis (Figure 5A). Gene Ontology (GO) analysis showed the DEGs in biological processes and cell components were involved in ER lipid metabolic process and ER lipid transport and binding (Figure 5A). Then we performed a heat map of DEGs associated with ER lipid metabolism, indicating the down-regulation of ER lipid synthesis-related genes, such as *SCD-1*, *ELOVL5*, *GPAT3*, *MOGAT1*, in the livers of HFD mice with TTR knockdown (Figure 5B). In addition, non-targeted lipidomics analysis revealed that the main different lipid species were TG and diacylglycerol (DAG) (Figure 5C). As the smooth ER is the primary site for hepatic DAG and TG production, our findings suggested that TTR knockdown was most likely to affect the glycerolipid synthesis at the ER level.

**Figure 5.**
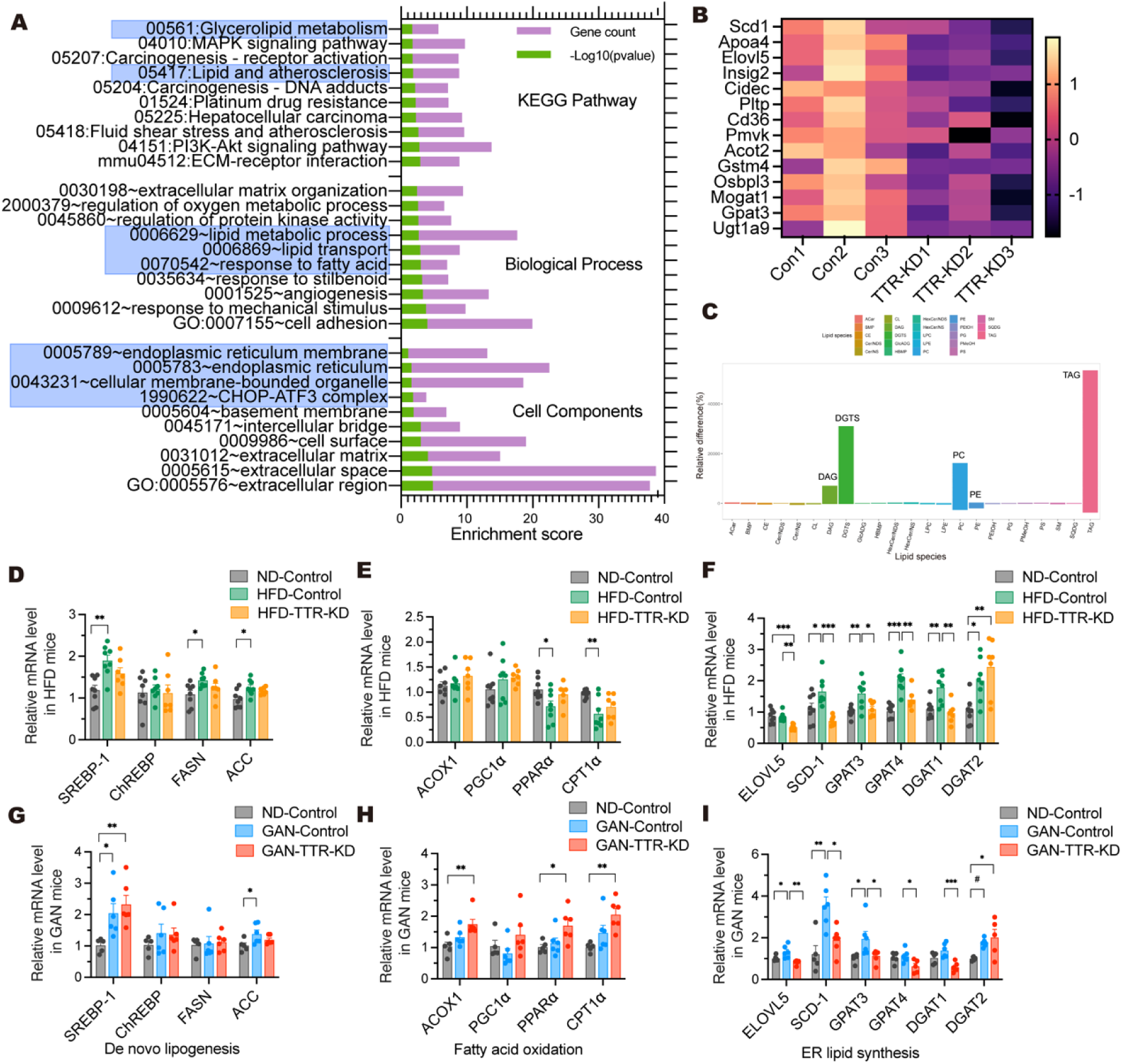
TTR knockdown downregulated the expression of ER lipid synthesis-related genes. (A-C) Bioinformatics analyses for the relationship between TTR knockdown and ER lipid metabolism in the liver of NAFLD mice (n = 3 per group). (A) KEGG pathway enrichment analysis and GO analysis for biological processes, cell components and molecular function regulated by differentially expressed genes (DEGs) (*P*< 0.05, ≥ 1.0-fold change). (B) Representative heat map of DEGs associated with ER lipid metabolism. (C) Non-targeted lipidomics analysis showed a lipid bar plot in differences of lipid species between TTR-KD mice and control mice fed on HFD. (D-I) qRT-PCR was used to confirm DEGs that were enriched in ER lipid metabolism-related pathways (n = 7-8 per group in HFD mice; n = 5-6 per group in GAN-diet mice). The mRNA expression levels of genes related to de novo lipogenesis (D, G), fatty acid oxidation (E, H) and ER lipid synthesis (F, I) in the livers of TTR-KD mice fed on HFD and GAN diet. Data are shown as the mean ± SEM. *P* values were from 1-way ANOVA. ^#^*P* = 0.07, \**P* < 0.05, \*\**P* < 0.01, \*\*\**P* < 0.001. KEGG, the Kyoto Encyclopedia of Genes and Genomes; GO, Gene Ontology.

Indeed, we confirmed that liver-specific TTR knockdown significantly down-regulated the expression of ER lipid synthesis-related genes (mainly *SCD-1*, *ELOVL5*, *GPAT3*, *GPAT4* and *DGAT1*) in the livers of HFD- or GAN diet-fed mice (Figure 5F, I). However, it didn’t alter the expression of genes related to de novo lipogenesis (DNL) or fatty acid oxidation (Figure 5D, E, G and H), indicating that TTR modulates lipid metabolism at the ER level.

### Liver-specific TTR knockdown inhibited hepatic ER stress in NAFLD mice, and upregulated SERCA2 expression both in vivo and in vitro

Prolonged and excessive ER stress plays a significant role in the development of NAFLD, through the exacerbation of hepatic steatosis, hepatocyte apoptosis, and inflammation(***Flessa et al., 2022; Lebeaupin et al., 2018***). We then assessed three ER stress pathway of PERK, IRE1α, and ATF6 in the livers of NAFLD mice fed on HFD and GAN diet. Compared with the control ND mice, HFD and GAN diet induced hepatic ER stress, as indicated by the increased protein levels of p-eIF2α, ATF6 and p-IRE1α (Figure 6A, B), all of which were reversed by TTR knockdown except for p-IRE1α (Figure 6A, B). We further observed the expressions of ER stress chaperone proteins, particularly GRP78, but did not detect any significant differences in the livers of HFD or GAN diet mice (Supplementary Figure 5).

**Figure 6.**
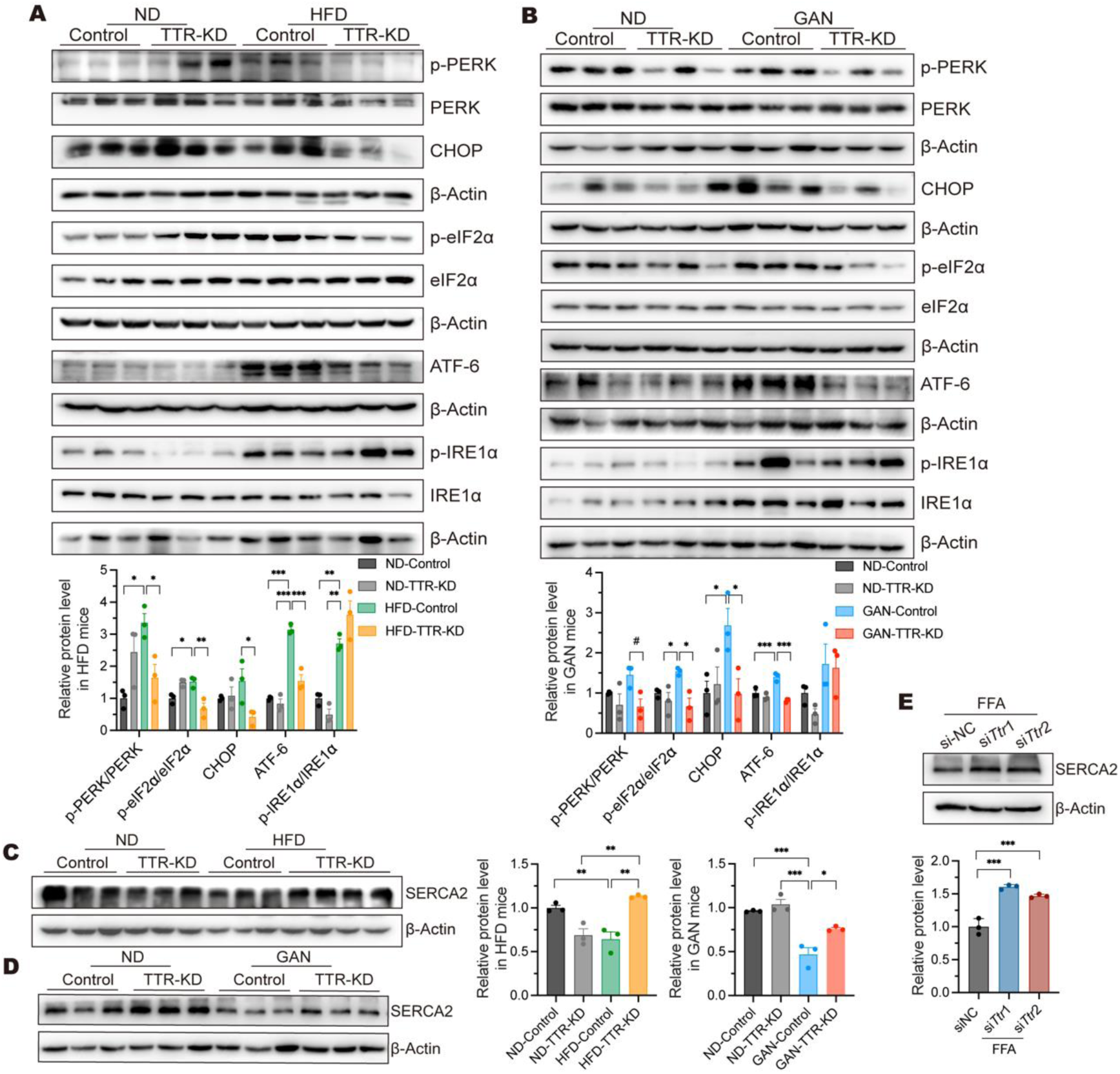
TTR knockdown inhibited ER stress and upregulated SERCA2 expression in the liver of NAFLD mice. Western blot analysis of the protein levels of p-PERK/p-eIF2α/CHOP and ATF6 signaling pathways in the liver of HFD mice (A) and GAN-diet mice (B). SERCA2 protein expression examined by western blot in the liver of HFD mice (C), GAN-diet mice (D), and FFA-induced AML12 cells transfected with siRNA (E). n = 7-8 per group in HFD mice; n = 5-6 per group in GAN-diet mice. Data are shown as the mean ± SEM. *P* values were from 1-way ANOVA. ^#^*P* = 0.06, \**P* < 0.05, \*\**P* < 0.01, \*\*\**P* < 0.001.

ER Ca^2+^ at high concentrations, primarily maintained by the SERCA, was essential for the ER function; and SERCA dysfunction can deplete ER Ca^2+^ content, impairing the activity of Ca^2+^-dependent folding enzymes and chaperones, thereby leading to ER stress(***Krebs et al., 2015***). SERCA2 is the predominant sub-type of SERCA in the liver. In our NAFLD mice fed on HFD or GAN diet, hepatic SERCA2 protein levels were significantly reduced compared to the control mice fed on ND, while TTR knockdown reversed inhibited SERCA2 levels (Figure 6C, D). Consistently, the upregulation of SERCA2 protein levels was found in steatotic AML12 cells transfected with *Ttr* siRNA (Figure 6E). These results suggest that TTR knockdown may increase SERCA2 levels and alleviate ER stress in the liver of NAFLD.

### TTR could be “taken up” by hepatocytes and localized close to the ER

TTR, synthesized in liver cells, is released into the circulation to function as a hormone transporter protein. Exogenous TTR enters various cells via internalization, including islet cells and skeletal muscle cell(***He et al., 2021; Dekki et al., 2012***). However, it remains unclear whether the same applies to the hepatocyte. We found that exogenous Alexa Fluor 488-labeled TTR probes emitted fluorescence signals within the cytoplasm of AML12 cells by continuous con-focal live-cell imaging. These signals overlapped with those of mCherry-ER-3, a fusion of mCherry fluorescent protein with the KDEL sequence, which serves as an ER marker (Figure 7A). In addition, we confirmed the co-localization of TTR and Calnexin, a marker protein for ER membrane by immunofluorescence staining in both AML12 and HepG2 cells (Figure 7B). All these results support the idea that secreted TTR could be taken up by hepatocytes and spatially localized near the ER.

**Figure 7.**
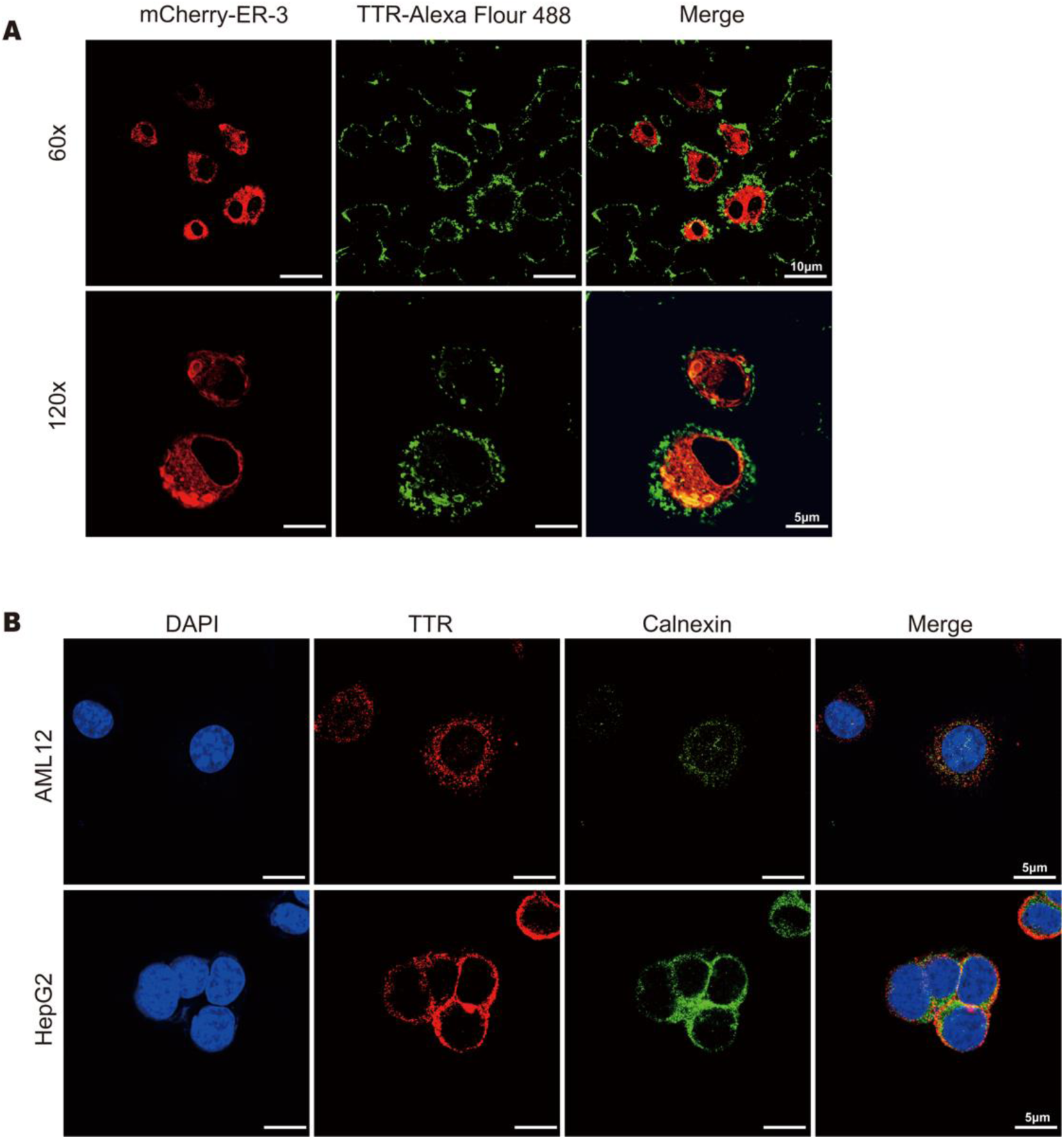
TTR could be “taken up” by hepatocytes and localized close to the ER. (A) AML12 cells transfected with the mCherry-ER-3 plasmid for 48 hours were then subjected to serum starvation for 6 hours, followed by the addition of 50 μg/mL Alexa Fluor 488 labeling TTR probes and incubated at 37°C in the dark for 30 minutes. After washing off the free probes, live cell imaging was immediately performed by confocal microscope. Alexa Fluor 488 labeling TTR was observed at an excitation wavelength of 488 nm, while the ER fluorescing red was visible at an excitation wavelength of 594 nm; scale bars: 10 μm (up), 5 μm (down). (B) Immunofluorescence staining of TTR and Calnexin in AML12 and HepG2 cells using confocal microscopy. TTR was observed at an excitation wavelength of 594 nm, and Calnexin at 488 nm. Nuclei were stained with DAPI. Scale bar: 5 μm.

### TTR silencing improved calcium homeostasis between the cytoplasm and the ER

As we have found that TTR’s knockdown in liver activated SERCA2 and inhibited ER stress, as well as the exogenous TTR uptake being closely localized to the ER, we further investigated the impact of TTR on the regulation of calcium homeostasis in hepatocytes. We found the cytosolic Ca^2+^ levels, using Fluo-8 AM incubation, were significantly increased after exogenous TTR treatment (10 μg/ml) over time (Figure 8A). While the ER Ca^2+^ levels, indicated by pcDNA-D1ER plasmid transfection, was markedly decreased (Figure 8B), suggesting potential impairment of SERCA2 function. Indeed, TTR overexpression combined with 5 μM thapsigargin (Tg, a SERCA inhibitor) pretreatment synergistically increased the fluorescence intensity of cytosolic Ca^2+^ compared to controls (Figure 8C); while TTR-KD stable cell lines showed a gentler increase in fluorescence intensity (Figure 8D). These findings suggest a possible role of TTR in the calcium homeostasis of hepatocytes by regulating SERCA2 activity.

**Figure 8.**
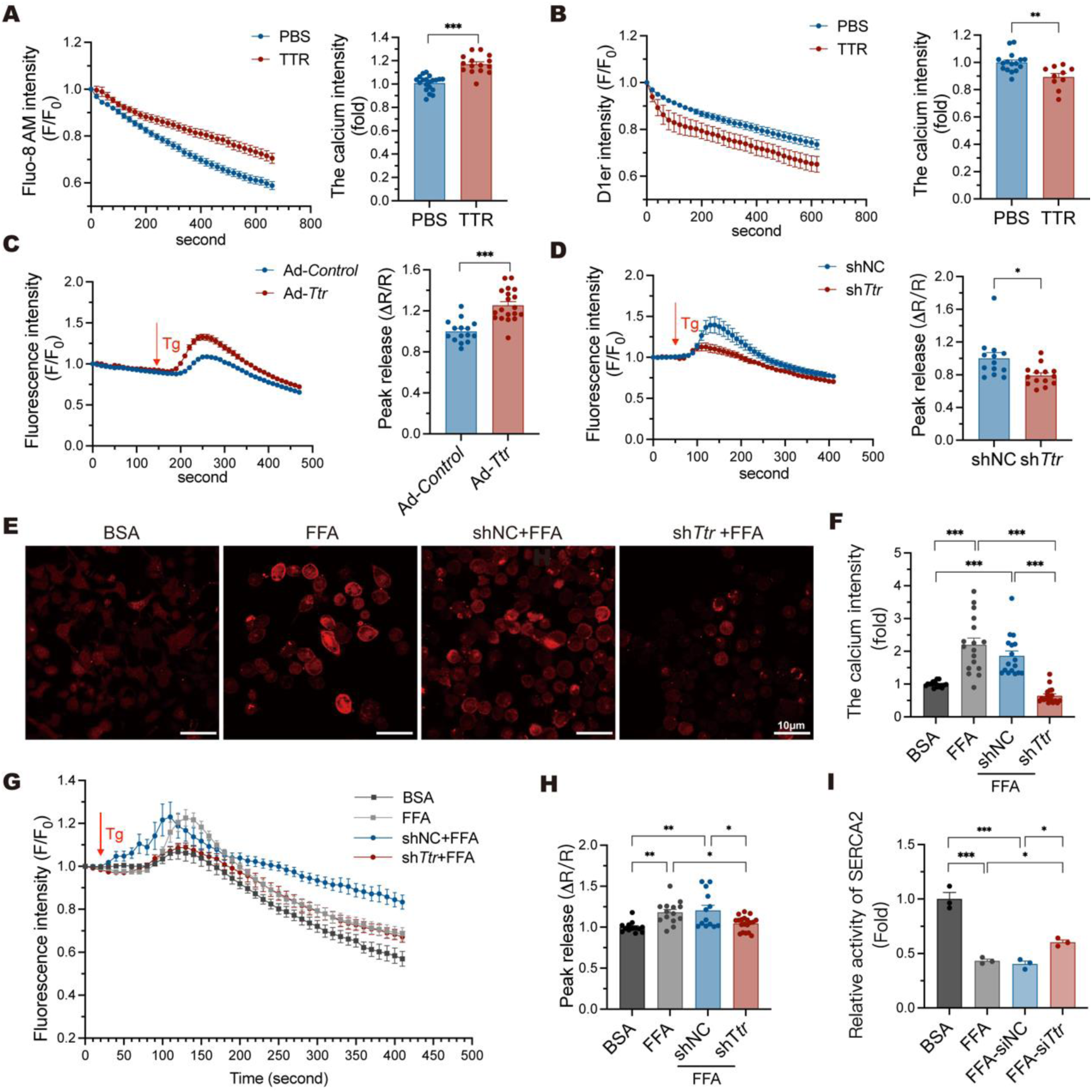
The effect of TTR on calcium homeostasis in AML12 cells. After the treatment of TTR (10 μg/ml) or PBS, the fluorescence intensity of Ca^2+^change was monitored in Fluo-8 AM-loaded AML12 cells (A) and in AML12 cells with pcDNA-D1ER plasmid transfection (B). 5 μM Tg was added to block calcium transport from the cytosol to the ER. The fluorescence intensity of Ca^2+^ change was detected in TTR-OE AML12 cells by adenovirus (C) and in TTR-KD stable cell lines (D). AML12 cells were incubated with BSA or FFA for 12 h. (E) Resting cytosolic calcium signals were detected in the TTR-KD stable cell lines by Cal-590 AM loading; scale bar: 10 μm. (F) Statistical chart of resting cytosolic calcium intensity in stable cell lines induced by FFA or BSA. (G) After the treatment of Tg (5 μM), the fluctuation of cytosolic calcium signal was monitored in Cal-590 AM-loaded AML12 cells or TTR-KD stable cell lines induced by FFA or BSA. (H) Statistical chart of the peak release ratio of cytosolic calcium intensity caused by Tg stimulation in stable cell lines induced by FFA or BSA. (I) Relative SERCA2 activity in BSA/FFA treated AML12 cells with *Ttr* siRNA infection or not. The activity of SERCA was calculated from the hydrolyzed ATP level (nmol) normalized with the protein content (mg) and reaction time (min). Data are shown as the mean ± SEM. *P* values were from 2-tail unpaired *t* test (A, B, C and D) and 1-way ANOVA (F, H). n = 10-20 cells per group. \**P* < 0.05, \*\**P* < 0.01, \*\*\**P* < 0.001. Tg, thapsigargin.

Abnormal calcium homeostasis contributes to NAFLD, we thus further assessed the calcium homeostasis as well as TTR’s effect on it in steatotic hepatocytes. FFA treatment significantly increased the basal cytosolic Ca^2+^ compared to the bovine serum albumin (BSA) treatment; while this increase was partially attenuated upon TTR knockdown (Figure 8E, F). Upon Tg stimulation, FFA-induced cytosolic Ca^2+^ fluctuation was exacerbated, and this effect was also flattened by TTR knockdown (Figure 8G, H). Compared to BSA-treated controls, FFA-treated AML12 cells exhibited significantly reduced SERCA2 activity, as measured by ATPase activity; while TTR knockdown reversed the SERCA2 activity inhibited by FFA (Figure 8I). Thus, in both steatotic and non-steatotic hepatocytes, TTR fulfilled a function similar to that of Tg, targeting SERCA2 activity.

### TTR co-localized with SERCA2 and interacted with SERCA2 in HEK293T cells and hepatocytes

Our prior findings suggested that the uptake of TTR led to its location in the ER area and might exert an inhibitory effect on SERCA2 activity; we then explored the interaction between TTR and SERCA2. We validated the co-localization of TTR and SERCA2, showing overlapping red and green fluorescence as yellow regions in AML12 and HepG2 cells using con-focal microscopy (Figure 9A). Then using co-immunoprecipitation (Co-IP), we confirmed direct binding between TTR and SERCA2 in HEK-293T cells co-transfected with TTR-FLAG and SERCA2-MYC plasmids (Figure 9B, C). In addition, the direct interaction between TTR and SERCA2 was also observed in AML12 cells transfected with TTR-FLAG plasmid (Figure 9D).

**Figure 9.**
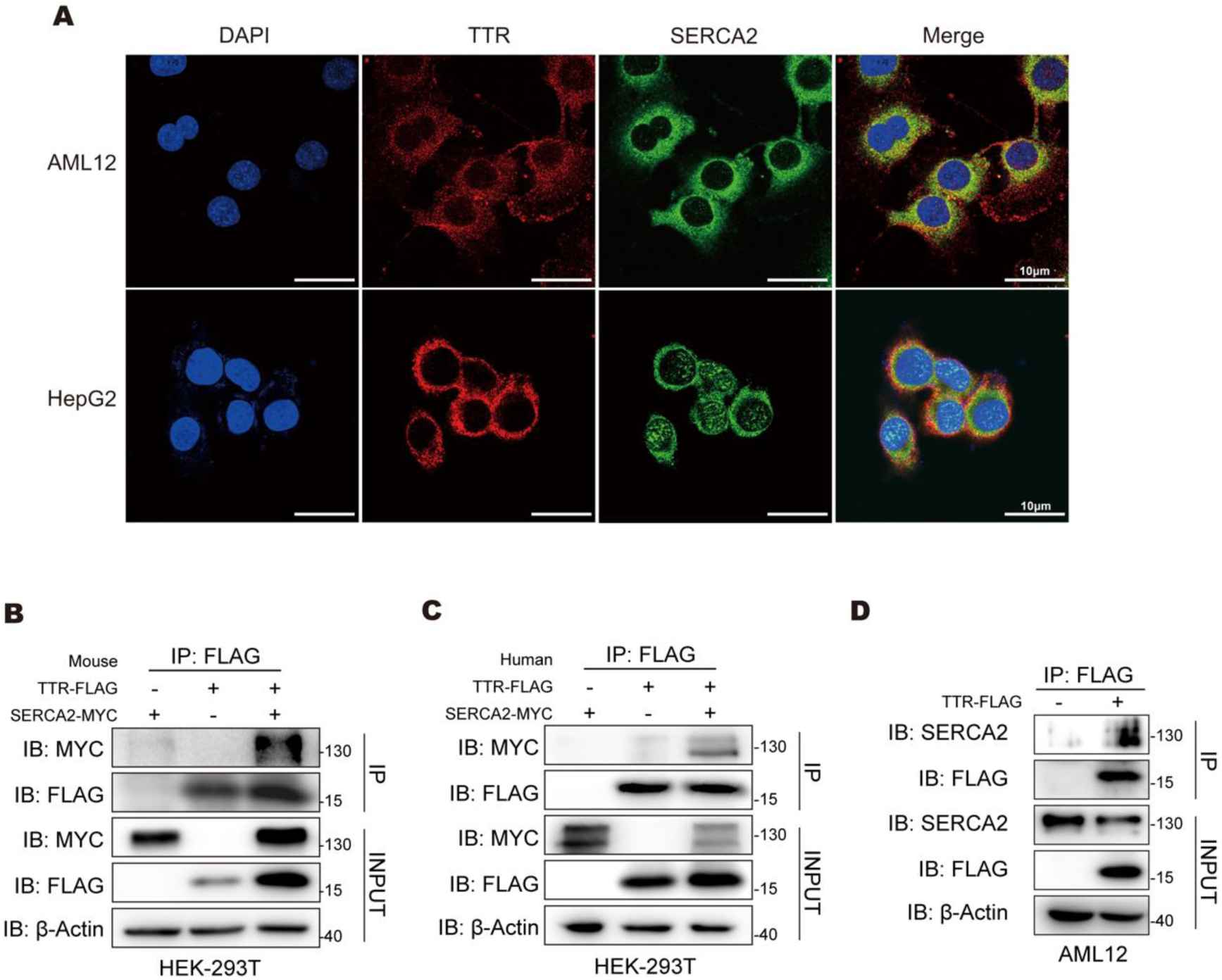
Protein interaction between TTR and SERCA2 in hepatocytes. (A) Immunofluorescence staining of TTR and SERCA2 in AML12 and HepG2 cells using confocal microscopy; scale bar: 10 μm. (B, C) HEK-293T cells overexpressed TTR-FLAG and SERCA2-MYC, and Co-IP was used to detect the protein interaction. (B) TTR and SERCA2 shared a common murine genetic origin. (C) TTR and SERCA2 shared a common human genetic origin. (D) AML12 cells overexpressed FLAG-TTR, and CoIP was used to detect the interaction between TTR-FLAG protein and endogenous SERCA2 protein. TTR-FLAG, TTR protein labeled with FLAG; SERCA2-MYC, SERCA2 protein labeled with MYC; Co-IP, co-immunoprecipitation.

## Conclusions

Elevated hepatic TTR expression and secretion contribute to the development of NAFLD under HFD feeding. Liver-specific TTR knockdown alleviates hepatic steatosis and inflammation in NAFLD mice, potentially by improving Ca^2+^ homeostasis through SERCA2 activation. This subsequently attenuates ER stress and reduces ER-related lipid synthesis, suggesting TTR as a potential therapeutic target for NAFLD prevention.

## Discussions

The pathogenesis of NAFLD is not yet fully understood, which poses a huge challenge to its treatment. Except for the THR-β agonist, Resmetirom, approved by FDA this year(***Kang et al., 2016***), there are no other FDA-approved drugs for the treatment of NAFLD. The accumulation of liver lipids, primarily TG, is the triggering factor, and inflammation is a crucial step in the progression of NAFLD. Thus, molecular factors that promote hepatic lipid deposition and inflammation may serve as potential targets for NAFLD prevention. To our knowledge, our study is the first to explore the role of TTR on NAFLD development; we found that liver-specific TTR knockdown improved calcium homeostasis via targeting SERCA2 activity, and reduced hepatic ER stress, lipid accumulation, and inflammation.

TTR is a homo-tetrameric protein primarily synthesized by both choroid plexuses of the brain and liver, serving as a transport carrier for thyroid hormones and retinol in the circulation. Recent studies have identified that TTR in the peripheral system is involved not only in the assessment of nutritional status but also associated with glucose metabolism and IR(***He et al., 2021; Zemany et al., 2015; Ingenbleek and Bernstein, 2015***). However, the effect of TTR on the pathogenesis and development of NAFLD remains unclear. We initially examined TTR levels in NAFLD state, and found a significant increase of *Ttr* mRNA expression in NASH patients compared to non-NAFLD controls. Then we observed that TTR overexpression exacerbated hepatic lipid deposition in HFD-fed mice and FFA-induced steatotic hepatocytes. In contrast, liver-specific TTR knockdown relieved lipid accumulation and inflammation in the livers of various NAFLD mouse models, indicating a role of TTR in the development of NAFLD.

Our prior studies have found that administration of purified TTR via the lateral ventricle inhibits appetite, contributing to body weight loss(***Zheng et al., 2016***); while in the periphery, high expression of TTR induces IR in ND mice(***He et al., 2021***). In addition, liver-specific TTR knockdown using a liver-targeting AAV vector, significantly improved IR in ob/ob mice but had no effect on body weight.

This suggests that TTR has distinct roles in both the central and peripheral systems(***He et al., 2021***). This may also explain why the global absence of TTR in mice does not improve metabolic phenotypes(***Marques et al., 2007; Rendenbach et al., 2013***). In our study, both GTT and ITT results showed that liver-specific TTR knockdown improved glucose tolerance and insulin sensitivity in NAFLD mice induced by HFD and GAN diet, independent of body weight changes, which supports a role of peripheral TTR in energy metabolism.

Consistently *in vivo* and *in vitro*, we observed that both HFD feeding and FFA treatment upregulated hepatic TTR expression. *This TTR induction may exacerbate IR, thereby accelerating hepatic lipid accumulation and serving as the initial hit in NAFLD development.* Through transcriptomic and lipidomic analyses and validation of key genes, we did find that TTR knockdown primarily inhibited lipid synthesis at the ER level, particularly the fatty acid elongation required for glycerolipid formation, whereas it had no effect on DNL and fatty acid oxidation. This is consistent with the spatial localization of exogenous TTR mainly in the ER after reuptake.

TTR synthesized by the liver may regulate hepatocyte metabolism through its intracellular actions and via a reuptake mechanism (i.e., autocrine action). The uptake of TTR occurs in several cell types(***Dekki et al., 2012*; *Divino and Schussler*, 1990; *Divino and Schussler*, 1990**). In this study, we validated TTR internalization in hepatocytes using TTR-fluorescent protein probes. However, the cell surface receptor mediating TTR uptake in hepatocytes requires further investigation.

ER stress is considered to be the second hit that drives the progression of NAFLD, following the initial lipid accumulation induced by IR(***Flessa et al., 2022*; *Lebeaupin et al., 2018***). Consistent with other studies, we confirmed increased ER stress in the livers of different NAFLD mouse models.

Sustained and excessive ER stress can, in turn, further amplify lipid accumulation, induce oxidative stress and inflammation(***Piccolis et al., 2019***). Calcium homeostasis disruption is a primary trigger of ER stress(***Krebs et al., 2015***). In NAFLD, the accumulation of lipids in hepatocytes inhibits Store-Operated Calcium Entry (SOCE) and the ER calcium pump SERCA2, while activating calcium efflux from the ER and its transfer to the mitochondria. This contributes to the impairment of the function of calcium-dependent molecular chaperones, and subsequently activating ER stress and mitochondrial dysfunction, accelerating the progression from steatosis to steatohepatitis(***Chen et al., 2021*; *Jacquemyn et al., 2017***). Multiple studies have shown impaired activity and reduced expression of liver SERCA2 in animal models of NAFLD(***Park et al., 2010*; *Kang et al., 2016***). We consistently found that SERCA2 protein levels were suppressed in NAFLD mice induced by HFD or GAN diet, as well as in steatotic AML12 cells. An increasing number of studies have confirmed that some small molecule compounds, including CDN1163(***Kang et al., 2016***), Maresin 1(***Jung et al., 2018***), and Matrine(***Gao et al., 2018***), targeting SERCA2 activity can delay the development of NAFLD.

We then found that TTR’s regulation of calcium homeostasis (increasing cytoplasmic calcium levels and reducing ER calcium stores) is very similar to the effect of the SERCA inhibitor, Tg. Further in *vitro* studies, we confirmed a direct interaction between TTR and SERCA2, and an inhibitory effect of TTR on SERCA2 activity. Thus, we propose that in the context of NAFLD, TTR may induce calcium homeostasis abnormalities by inhibiting SERCA2 activity, thereby promoting ER stress and lipid synthesis. With growing recognition of the link between calcium homeostasis and NAFLD pathogenesis, SERCA2 activity modulators have emerged as promising therapeutic targets for NAFLD drug development. We confirmed in this study that TTR regulates SERCA2 activity, but the specific molecular mechanisms by which TTR directly inhibits SERCA2 activity warrant further research.

In summary, we establish that TTR knockdown alleviates NAFLD by restoring hepatocyte calcium homeostasis via SERCA2 upregulation, thereby attenuating ER stress and suppressing ER-dependent lipid synthesis in the liver. These findings further support the crucial role of calcium signaling in NAFLD and offer new insights for the development of novel calcium channel-targeted therapeutic approaches for NAFLD, e.g., targeting TTR.

## Materials and methods

### Sex as a biological variable

Our study examined male mice because male animals exhibited less variability in phenotype. It is unknown whether the findings are relevant for female mice. Therefore, we clearly stated which sexes were analyzed in the respective experiments.

### Liver specimens from patients with NAFLD

We collected liver sample sections from Sir Run Run Shaw Hospital, Zhejiang University. Liver samples of 5 male patients diagnosed with NASH through liver biopsy in the Infectious Disease Department were selected. Another 5 male patients who underwent cholecystectomy in the General Surgery Department and were excluded from fatty liver by ultrasound were selected as non-NAFLD controls. The liver tissue sections were analyzed independently by senior pathologists according to the SAF scoring system (“S” standing for steatosis was scored from 0 to 3, “A” standing for ballooning degeneration and lobular inflammation was scored from 0 to 4, “F” standing for fibrosis was scored from 0 to 4). The general clinical information of 10 selected patients was collected (Supplementary Table 1).

### Animals

Male C57BL/6 mice (aged 5-8 weeks) were obtained from Shanghai SLAC Laboratory Animal Co., Ltd., housed in a temperature-controlled environment with 12-hour light/12-hour dark cycle and received food and water ad libitum. Mice were fed on a ND (MD17131, Medicience), a HFD (M101160, Moldiets) or a GAN diet (D09100310, Research Diet), respectively. Body weight was measured weekly. After treatments, the mice were sacrificed, and the plasma and liver tissues were collected for analysis.

Cohort 1: Male C57BL/6J mice were randomly divided into four groups (n = 4 each): ND-Control group, ND-TTR overexpression group (ND-TTR-OE), HFD-Control group, and HFD-TTR-OE group. The mice were fed with ND or HFD for 19 weeks. On weeks 9 and 11, each mouse received a tail vein injection of adenovirus (1 × 10^9^ plaque-forming units/mouse; GeneChem), with the control groups receiving control adenovirus (Ad-Control) and the TTR-OE groups receiving Ad-*Ttr*.

Cohort 2: Male C57BL/6J mice were randomly divided into four groups: ND-Control group (n = 8), ND-TTR knockdown group (ND-TTR-KD) (n = 8), HFD-Control group (n = 8), and HFD-TTR-KD group (n = 7). The mice were fed with ND or HFD for 19 weeks. On weeks 9 and 11, each mouse received a tail vein injection of recombinant adeno-associated virus bearing TBG promoter (1 × 10^11^ plaque-forming units/mouse; GeneChem), with the control groups receiving control adeno-associated virus (AAV-TBG-Control) and the TTR-KD groups receiving AAV-TBG-sh*Ttr*.

Cohort 3: Male C57BL/6J mice were randomly divided into four groups (ND group n = 5, GAN group n = 6): ND-Control group, ND-TTR-KD group, GAN-Control group, and GAN-TTR-KD group. The mice were fed with ND or GAN diets for 17 weeks. On weeks 1 and 9, each mouse received a tail vein injection of AAV bearing TBG promoter, with the control groups receiving AAV-TBG-Control and the TTR-KD groups receiving AAV-TBG-sh*Ttr*.

### Cell culture and treatment

AML12 cells were purchased from the American Type Culture Collection (ATCC) and grown in Dulbecco’s Modified Eagle Medium/Nutrient Mixture F-12

(DMEM/F12) supplemented with 10% fetal bovine serum, 0.1 μM dexamethasone (D4902; Sigma-Aldrich), insulin-transferrin-selenium (I3146; Sigma-Aldrich) and 1% penicillin-streptomycin. HepG2 and HEK293T cells were purchased from ATCC and cultured in DMEM with 10% fetal bovine serum and 1% penicillin-streptomycin at 37°C with 5% CO2.

To induce steatosis, the cells were treated with 0.75 mM FFA (palmitic acid: oleic acid, 1:2; Sigma-Aldrich) for 12 hours at 37°C. For TTR overexpression, AML12 cells were infected with 1.25×10^7^ plaque-forming units/mL (PFU/mL) of Ad-Control or 5×10^7^ PFU/mL Ad-*Ttr* for 48 hours. For TTR knockdown, AML12 cells were treated with small interfering (si)-*Ttr*1 (5’-GUCCU CUGAU GGUCA AAGUT T-3’), si-*Ttr*2 (5’-CCACA GAUGA GAAGU UUGUT T-3’) and si-NC (5’-UUCUC CGAAC GUGUC ACGUT T-3’) for 48 hours, which were synthesized by GenePharm. AML12 cells were transfected using Rfect siRNA transfection reagent (Cat#:11013, Baidai Biotechnology, China) according to the manufacturer’s protocol.

For achieving TTR-KD stable cell lines, shRNA lentiviral particle was synthesized and transfected.

First, lentiviral constructs for shRNA targeting TTR (sh*Ttr*) and shRNA control were purchased from Tsingke Biotechnology. The lentiviral sh*Ttr* constructs were designed with the following target sequences: 5’-AACAGTGTTCTTGCTCTATAA-3’ and 5’-CCACAGATGAGGAAGTTTGT-3’. Then, lentiviruses were produced by transfection of HEK293T cells with lentiviral constructs and pMD2.G and psPAX2 packaging constructs (Addgene). PolyJet reagents (Signagen) were used for transfection and the supernatants containing the lentiviruses were centrifuged and filtered, and then stored at -80 °C. We incubated AML12 cells with thawed lentivirus-sh*Ttr* or control lentivirus, together with polybrene (GeneChem) for 48 hours. For the selection of cells stably transfected with shRNA, the medium was replaced with a 10% serum medium containing 3 μg/mL puromycin (Solarbio, China) and incubated for 48 to 72 hours. Cells were replenished with medium every 3 to 4 days until several puromycin-resistant colonies were identified. Efficacy and successful shRNA transduction were verified by qRT-PCR.

### Localization of Alexa Fluor 488 labeling TTR in AML12 cells

Alexa Fluor 488 labeling TTR probes were synthesized as described previously(***He et al., 2021***). The mCherry-ER-3 plasmid (Addgene) was a gift from Prof. Yingke Xu (Zhejiang University). AML12 cells transfected with the mCherry-ER-3 plasmid for 48 hours were then subjected to serum starvation for 6 hours, followed by the addition of 50 μg/mL Alexa Fluor 488 labeling TTR probes and incubated at 37°C in the dark for 30 minutes. The cells were then washed with PBS to completely remove the free probes, and live cell imaging was immediately performed by confocal microscope. Alexa Fluor 488 labeling TTR was observed at an excitation wavelength of 488 nm, while the ER fluorescing red was visible under an excitation wavelength of 594 nm.

### Confocal microscopy for Ca^2+^ imaging

For measuring the cytosolic Ca^2+^ concentration in AML12 cells, cells were incubated with 2 μM Fluo-8 AM (142773, Abcam) in Hank’s Balanced Salt Solution (HBSS) without Ca^2+^ and Mg^2+^ for 30 min at 37°C in the dark. For measuring the Ca^2+^ in the ER, cells were transfected with pcDNA-D1ER plasmid (36325, Addgene) for 48 hours, which was expressed to be D1ER calcium indicator equipped with an ER-targeting sequence and fluorescent proteins. For measuring the cytosolic Ca^2+^ levels in stable cell lines with TTR knockdown, cells were incubated with 2 μM Cal-590 AM (20510, AAT Bioquest) in calcium-free HBSS for 90 min at 37°C in the dark. 5 μM thapsigargin (Sigma) was used to stimulate Ca^2+^ release. Once added 10 μg/mL TTR or PBS or 5 μM Tg, cells were imaged every 10 to 20 seconds for 6-10 min using confocal microscopy at an excitation wavelength of 488 nm. The calcium signal was presented as the mean of the relative change in fluorescence intensity normalized to baseline intensity (ΔF/F) in 10-20 AML12 cells.

### SERCA activity analysis

To facilitate the transport of calcium from the cytosol into the ER lumen, SERCA facilitates the hydrolysis of ATP into ADP and inorganic phosphate. As a result, the level of inorganic phosphate serves as an indicator of SERCA activity. SERCA activity was assessed using an ultra-micro Ca²⁺-ATPase assay kit (GMS50241, GENMED). After trypsinization and centrifugation, AML12 cells were suspended in normal saline and disrupted via sonication. The protein concentration was quantified using the BCA method.

### Immunofluorescence staining

After 20 minutes of immersion in PBS containing 0.2% Triton X-100, the cell samples were fixed with 4% paraformaldehyde for 20 minutes, then blocked with 5% BSA for 1 hour and incubated with the primary antibody at 4°C overnight before being incubated with fluorescent-dye–conjugated secondary antibodies (Alexa Fluor 488/594, Invitrogen; 1:500). Primary antibodies used: anti-mouse-TTR (204997, Abcam; 1:100), anti-rabbit-SERCA2 (3625, Abcam; 1µg/mL), anti-rabbit-Calnexin (2670, CST; 1:100). Cell nuclei were stained with DAPI. Images were acquired by a confocal microscope.

### Co-immunoprecipitation

CMV-FLAG-TTR (mouse) plasmid, pcDNA3.1-FLAG-TTR (human) plasmid, pCMV-MYC-SERCA2 (mouse) plasmid, and CMV-MYC-SECRA2 (human) plasmid were purchased from GeneChem. For TTR-FLAG and SERCA2-MYC co-immunoprecipitation, HEK293T cells were transfected with mouse or human TTR-FLAG and SERCA2-MYC plasmids for 48 hours. For TTR-FLAG and SERCA2 co-immunoprecipitation, TTR-FLAG were overexpressed in AML12 cells. The cell lysates were centrifuged at 12,000 g for 15 min at 4°C. The supernatants were further incubated with primary antibodies against FLAG or MYC or IgG at 4°C overnight, then precipitated with Protein A/G beads (sc-2003, Santa Cruz) at 4°C for 4 hours. The beads were washed five then boiled in SDS buffer. SERCA2-MYC, TTR-FLAG and SERCA2 protein levels were determined via western blot.

Please refer to the Supplemental Methods for detailed materials and methods.

### Statistics

GraphPad Prism 9 and image J were used to analyze the values in this study. All data are presented as mean ± SEM, unless otherwise indicated and analyzed by unpaired 2-tailed t test or 1-way ANOVA if more than 2 groups were compared. Statistical significance was set at *P* < 0.05.

### Study approval

All protocols and animal studies were performed in accordance with the Guide for the Care and Use of Laboratory Animals by the US NIH (NIH Publication No. 85-23; National Academies Press, 2011) and approved by the Animal Ethics Committee of Zhejiang Chinese Medical University (Approval No. 20190429-04). The human study conforms to the ethical guidelines of the 1975 Declaration of Helsinki as reflected in a priori approval by the Ethics Committee of Sir Run Run Shaw Hospital. All human samples were acquired ethically with patient/guardian consent following institutional guidelines.

## Data Availability

Values for all data points in graphs are reported in the Supporting Data Values file. For more details on the material and methods used, please contact the corresponding author.

## Acknowledgements

We want to thank all participants for taking part in our study and consenting to donate extra Liver samples for research use. We thank Prof. Yingke Xu (Zhejiang University) for kindly providing the valuable plasmid (mCherry-ER-3) and for excellent technical assistance. This work was supported by the National Natural Science Foundation of China [81870583, 2018] and Zhejiang Provincial Natural Science Foundation of China [ZCLQN25H0701, 2025]. The graphical abstract was created with BioRender.com and were approved for publication. The contents of the published material are solely the responsibility of the individual authors and do not reflect the view of any of the funding bodies.

## Notes

### Competing Interest Statement

The authors have declared no competing interest.

### Author Declarations

Animal studies were approved by the Animal Ethics Committee of Zhejiang Chinese Medical University (Approval No. 20190429-04). The human study conforms to the ethical guidelines of the 1975 Declaration of Helsinki as reflected in a priori approval by the Ethics Committee of Sir Run Run Shaw Hospital.

